# Reporting right ventricular strain and the RV/LV diameter ratio in 1,534 CT pulmonary angiography reports of acute pulmonary embolism

**DOI:** 10.64898/2026.11.27.26368979

**Authors:** Jatin Naidu, Vasanth Baskaradoss

**Author notes:** Corresponding author: Dr Jatin Naidu, Queen Elizabeth Hospital, Stadium Road, London SE18 4QH.

## Abstract

**Purpose:** To quantify how CT pulmonary angiography (CTPA) reports documented right ventricular (RV) strain and the RV-to-left ventricular (LV) diameter ratio in acute pulmonary embolism (PE) before the 2026 multisociety reporting recommendation.

**Methods:** This retrospective cross-sectional study included 1,534 CTPA reports from MIMIC-IV-Ext-PE (2008–2022), one per patient. Rule-based extraction classified overall strain statements and identified four CT features. A physician reviewed all 22 reports flagged for RV/LV ratio wording and independently coded a random held-out sample of 100 reports. Multivariable logistic regression, including approximate examination period, assessed factors associated with documentation gaps.

**Results:** Five reports (0.3%, 95% confidence interval [CI] 0.1-0.8%) gave an exact RV/LV ratio, and seven (0.5%) gave a threshold or range. Of 412 reports (26.9%) without an overall strain statement, 39 documented at least one coded CT feature. In total, 373 reports (24.3%, 95% CI 22.2-26.5%) documented neither. Isolated subsegmental PE was associated with higher adjusted odds of lacking an overall statement (odds ratio 2.76, 95% CI 2.02-3.78). These odds declined over time (2020–2022 vs 2008–2010: 0.14, 95% CI 0.06-0.31), and the association with a structured organ-system heading attenuated after this adjustment (0.99, 95% CI 0.69-1.43). Agreement with manual physician coding in classifying overall strain statements was 93.0% (Cohen’s κ 0.89, 95% CI 0.81-0.97).

**Conclusion:** Exact RV/LV ratios were rarely documented, and nearly one quarter of reports contained neither an overall strain statement nor any of the four CT features. Omission declined over time. These estimates provide a single-center baseline for evaluating uptake of the 2026 recommendation.

## Introduction

Computed tomography pulmonary angiography (CTPA) establishes the diagnosis of acute pulmonary embolism (PE) and can demonstrate right ventricular (RV) enlargement on the same examination. The ratio of RV to left ventricular (LV) diameter contributes to early risk stratification alongside clinical status and cardiac biomarkers, and the 2019 European Society of Cardiology (ESC) guideline already listed an RV/LV ratio of 1.0 or more on CTPA as a sign of RV dysfunction [1]. An elevated RV/LV ratio on CTPA has been associated with all-cause and PE-related mortality [2]. The ratio was therefore clinically relevant before reporting guidance addressed it directly. The 2025 ESC and Fleischner Society consensus identified the RV/LV ratio as a key reporting element, and the 2026 AHA/ACC multisociety guideline recommends reporting the measured numerical ratio in acute PE [3,4].

An overall interpretation of RV strain, a description of individual CT features, and a measured RV/LV ratio convey distinct information. For example, septal flattening may be described without an overall interpretation, while a statement that strain is absent may provide no ventricular measurement. Previous audits documented variability in CTPA reporting [5,6]. In a single-center audit of 50 positive CTPA reports, 19 contained an explicit statement about right heart strain. That audit grouped RV size and RV/LV ratio within a single reporting item, leaving the frequency of exact numerical reporting unresolved [6].

We examined 1,534 positive CTPA reports. We distinguished exact ratios from numerical thresholds or ranges and qualitative descriptions, and identified reports in which individual CT features were documented without an overall strain statement. We also examined whether documentation varied over time and by report format, emergency department (ED) linkage, and PE location. These analyses provide a benchmark, preceding the 2025 and 2026 reporting recommendations, for auditing both numerical ratio reporting and the information conveyed about RV strain.

## Materials and methods

### Study design and cohort

We conducted a retrospective cross-sectional analysis of CTPA reports from MIMIC-IV-Ext-PE [7,8], which was derived from MIMIC-IV v3.1 at Beth Israel Deaconess Medical Center, Boston [9,10], with report text from MIMIC-IV-Note [11]; all three datasets are distributed through PhysioNet [12]. The source database covers 2008–2022. MIMIC shifts dates for de-identification, so exact examination years were unavailable; we estimated them as described under Outcomes and statistical analysis. MIMIC-IV-Ext-PE contains 19,942 CTPA reports from 15,875 patients [8]. Two physicians labeled each report as acute PE, subsegmental PE (acute PE confined to the subsegmental arteries), chronic PE, equivocal, or negative after reviewing extracted PE-related sentences and, where needed, full reports. We included the 1,591 reports labeled acute or subsegmental PE (1,358 and 233, respectively) and excluded the 18,351 reports labeled negative (17,902), chronic PE (345), or equivocal (104). Retaining the first dated positive report for each patient excluded 57 later positive reports from patients already included (49 acute and 8 subsegmental), leaving 1,534 reports: 1,309 with PE extending proximal to the subsegmental arteries and 225 with isolated subsegmental PE. Reporting follows the STROBE and RECORD statements [13,14].

### Report coding and verification

We applied Python regular expressions to full report text after collapsing line breaks and repeated spaces. The rules classified overall statements about right heart or RV strain, dysfunction, or overload as present, hedged, absent, or not stated. Requests for assessment in the clinical indication were excluded. Explicit negation was coded as absent. Qualifying language or a recommendation for echocardiography to assess named RV strain was coded as hedged. Unqualified assertions were coded as present. When categories co-occurred, present took precedence over hedged, followed by absent.

We recorded whether the impression contained an overall strain statement and whether the report documented any of four coded CT features: RV/LV size or ratio information, RV size, interventricular septal configuration, or contrast reflux into the inferior vena cava or hepatic veins. A feature was coded whenever the report described it, whether as normal or abnormal; for example, “normal RV size” and “no septal flattening” were both coded as documentation. Septal coding excluded pulmonary interlobular septa. We categorized RV/LV documentation as an exact value, a numerical threshold or range, a ratio named without a number, paired diameters without a ratio, or a qualitative comparison alone.

To identify numerical ratios the coding rules might have missed, we ran a supplementary search of all 1,534 reports for ratio terms near digits, colon notation, and RV or LV diameter measurements. A resident physician reviewed the full text of all 22 reports flagged by the coding rules or the supplementary search. The confirmed categories, including the paired-diameter report found only by the supplementary search, were incorporated into the final classifications used in Tables 2-4.

**Table 1.** Cohort characteristics.

| **Characteristic** | **Value** |
|---|---|
| Estimated age at CTPA, median (IQR), years | 64 (52–75) |
| Women | 832 (54.2%) |
| Isolated subsegmental PE | 225 (14.7%) |
| CTPA during an ED stay | 632 (41.2%) |
| Structured organ-system heading (report-format proxy) | 550 (35.9%) |
| Estimated examination period 2008–2010 | 224 (14.6%) |
| Estimated examination period 2011–2013 | 203 (13.2%) |
| Estimated examination period 2014–2016 | 418 (27.2%) |
| Estimated examination period 2017–2019 | 621 (40.5%) |
| Estimated examination period 2020–2022 | 68 (4.4%) |

Values are n (%) unless indicated. CTPA, computed tomography pulmonary angiography; ED, emergency department; IQR, interquartile range; PE, pulmonary embolism.

**Table 2.**
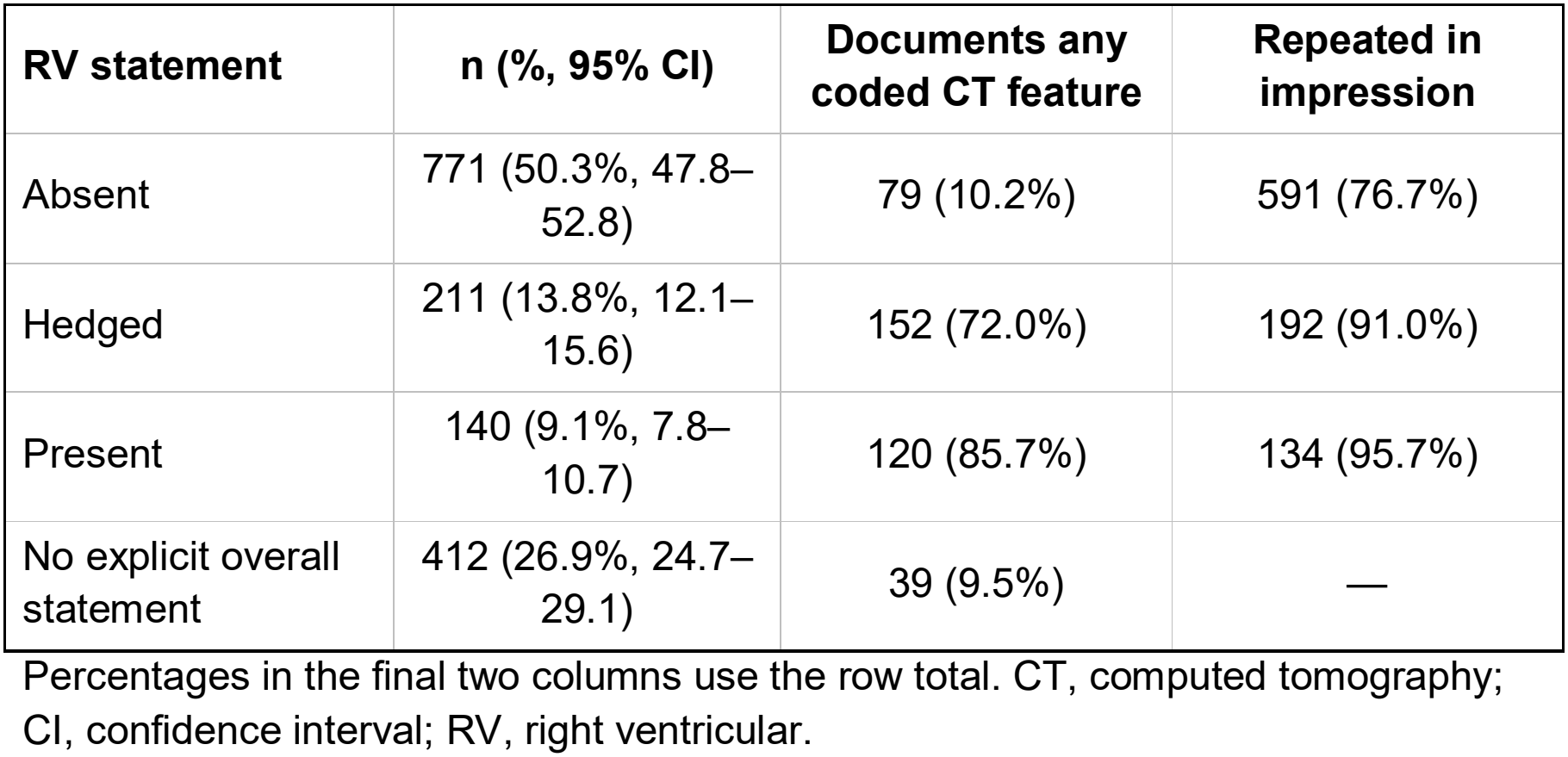
Overall RV strain statements, coded CT features, and impression documentation.

### Rule development and held-out evaluation

The resident physician manually coded 150 development reports while blinded to the automated classifications. These comprised 100 randomly selected reports and 50 enriched for strain statements and coded CT features. We revised the rules after reviewing discrepancies, then compared the revised classifications with blinded manual coding of 100 randomly selected reports outside the development sample. A consultant radiologist advised on general interpretation queries. The resident physician coded both samples and made all final coding decisions. No AI tool was used for reference coding.

### Outcomes and statistical analysis

The primary documentation outcome was omission of an overall RV strain statement; the secondary outcome was omission of both an overall statement and all four CT features. We fitted a multivariable logistic regression model for each outcome, adjusting for age, sex, PE location, ED linkage, report format, and approximate examination period. Age at CTPA was estimated as the MIMIC anchor age plus the interval between the report year and the anchor year. ED linkage indicated that CTPA occurred during a linked ED stay. The presence of the heading “HEART AND VASCULATURE” served as a proxy for structured report format. Because the heading may reflect when a report was written as well as its format, the models included examination period. MIMIC shifts dates while preserving intervals, so we estimated each examination year as the midpoint of the patient’s three-year anchor year group plus the difference between the shifted report year and the shifted anchor year (accurate to within 1 year), and grouped the estimates into consecutive three-year periods (2008–2010 to 2020–2022), with 2008–2010 as the reference. To show the effect of this adjustment, we also fitted the models without examination period. In sensitivity analyses, we replaced age at CTPA with the MIMIC anchor age and, separately, reclassified reports as having no overall statement when their only indication of strain was a recommendation for echocardiography to assess it.

We calculated Wilson 95% confidence intervals (CIs) for proportions and used 2,000 bootstrap resamples to estimate CIs for Cohen’s κ. Analyses were performed in Python 3.13 using statsmodels.

## Results

### Cohort and RV documentation

The cohort included 1,534 patients (median estimated age at CTPA 64 years, interquartile range 52–75 years). There were 832 women (54.2%) and 632 ED-linked examinations (41.2%) (Table 1). An overall RV strain statement was absent from 412 reports (26.9%). Of these, 39 documented at least one coded CT feature, leaving 373 reports (24.3%) with neither an overall statement nor any coded CT feature. A coded CT feature was documented in 79 of 771 reports (10.2%) stating that strain was absent and in 120 of 140 (85.7%) stating that strain was present (Table 2).

The coding rules flagged 21 reports with ratio-related wording, comprising 14 that used a ratio term and seven that provided a qualitative comparison alone. The supplementary search identified one report giving paired diameters without a stated ratio and found no further numerical ratios. Among the 14 reports using a ratio term, five reported an exact value, seven a threshold or range, and two gave no number (Table 3). Of the 12 reports with an exact value, threshold, or range, seven stated strain was present, three hedged, one stated strain was absent, and one provided no overall statement.

**Table 3.** RV/LV ratio documentation and other coded CT features.

| **Documentation** | **n (%, 95% CI)** |
|---|---|
| RV/LV ratio: exact numerical value | 5 (0.3%, 0.1–0.8) |
| RV/LV ratio: numerical threshold or range | 7 (0.5%, 0.2–0.9) |
| Any numerical RV/LV ratio | 12 (0.8%, 0.4–1.4) |
| Ratio named without a number | 2 (0.1%, 0.0–0.5) |
| RV and LV diameters given, ratio not stated | 1 (0.1%, 0.0–0.4) |
| Qualitative RV–LV size comparison only | 7 (0.5%, 0.2–0.9) |
| Interventricular septal configuration | 310 (20.2%, 18.3–22.3) |
| RV size described | 161 (10.5%, 9.1–12.1) |
| Contrast reflux into IVC or hepatic veins | 77 (5.0%, 4.0–6.2) |
| Any coded CT feature | 390 (25.4%, 23.3–27.7) |

The five RV/LV categories are mutually exclusive. “Any numerical RV/LV ratio” is the subtotal for exact values and thresholds or ranges. CI, confidence interval; IVC, inferior vena cava; RV, right ventricle; LV, left ventricle.

### ED-linked examinations

Among the 632 ED-linked examinations, two reports (0.3%, 95% CI 0.1-1.1%) gave an exact RV/LV ratio. None gave a threshold or range, a ratio without a number, or paired diameters, and two gave a qualitative comparison alone. An overall RV strain statement was absent from 106 ED-linked reports (16.8%, 95% CI 14.1-19.9%). Of these, 11 documented at least one coded CT feature, leaving 95 reports (15.0%, 95% CI 12.5-18.0%) with neither an overall statement nor any coded CT feature. Strain was stated as absent in 348 ED-linked reports (55.1%), hedged in 113 (17.9%), and present in 65 (10.3%); 26 of the 348 reports stating that strain was absent (7.5%) documented a coded CT feature. Among the 902 examinations without ED linkage, three reports (0.3%) gave an exact ratio and seven (0.8%) a threshold or range; 306 (33.9%) lacked an overall statement, and 278 (30.8%) documented neither an overall statement nor any coded CT feature.

### Factors associated with documentation gaps

Omission of an overall statement declined across examination periods, from 56.7% (127/224) in 2008–2010 to 13.2% (9/68) in 2020–2022 (Table 4). The structured heading appeared in no report from 2008–2013, in 70 of 418 (16.7%) from 2014–2016, and in 480 of 689 (69.7%) from 2017–2022. In the adjusted model (n = 1,534; 412 reports without an overall statement), the odds of omission were lower in each later period than in 2008– 2010 (2020–2022: adjusted odds ratio [OR] 0.14, 95% CI 0.06-0.31). The odds were higher for isolated subsegmental PE (adjusted OR 2.76, 95% CI 2.02-3.78) and lower for ED-linked examinations (adjusted OR 0.59, 95% CI 0.44-0.79). The association with a structured heading attenuated after adjustment for examination period (adjusted OR 0.99, 95% CI 0.69-1.43, compared with 0.53, 95% CI 0.40-0.70, without this adjustment). The secondary-outcome model gave similar estimates (isolated subsegmental PE, adjusted OR 3.04, 95% CI 2.21-4.17; ED linkage, 0.62, 0.46-0.84; structured heading, 1.03, 0.70-1.53; 2020–2022 vs 2008–2010, 0.14, 0.06-0.32). Using anchor age instead of age at CTPA gave near-identical estimates (isolated subsegmental PE, 2.77, 95% CI 2.03-3.78; ED linkage, 0.59, 0.44-0.79; structured heading, 0.99, 0.69-1.43; 2020–2022 vs 2008– 2010, 0.13, 0.06-0.30). Reclassifying five reports whose only qualifier was an echocardiography recommendation reduced the hedged proportion from 13.8% to 13.4% and increased the proportion with no overall statement from 26.9% to 27.2%.

**Table 4.** Factors associated with omission of an overall RV strain statement.

| **Factor** | **No overall statement, %** | **Adjusted OR (95% CI)** |
|---|---|---|
| Structured heading vs none | 17.5 vs 32.1 | 0.99 (0.69–1.43) |
| Isolated subsegmental vs more proximal PE | 44.4 vs 23.8 | 2.76 (2.02–3.78) |
| CTPA during ED stay vs not | 16.8 vs 33.9 | 0.59 (0.44–0.79) |
| Women vs men | — | 0.81 (0.64–1.04) |
| Estimated age at CTPA, per year | — | 1.00 (0.99–1.00) |
| Examination period 2011–2013 vs 2008–2010 | 34.0 vs 56.7 | 0.45 (0.30–0.67) |
| Examination period 2014–2016 vs 2008–2010 | 23.2 vs 56.7 | 0.28 (0.19–0.41) |
| Examination period 2017–2019 vs 2008–2010 | 17.7 vs 56.7 | 0.20 (0.13–0.31) |
| Examination period 2020–2022 vs 2008–2010 | 13.2 vs 56.7 | 0.14 (0.06–0.31) |

The model includes all covariates shown. Examination period was estimated from MIMIC anchor year groups (accurate to within 1 year). Without examination period in the model, adjusted ORs were 0.53 (0.40–0.70) for a structured heading, 2.64 (1.95–3.58) for isolated subsegmental PE, and 0.46 (0.35–0.60) for ED linkage. Age at CTPA was estimated as the MIMIC anchor age plus the interval between the report year and the anchor year; models using anchor age gave near-identical estimates (see text). Estimates for the secondary outcome (neither an overall statement nor any coded CT feature) are reported in the text. CI, confidence interval; CT, computed tomography; CTPA, computed tomography pulmonary angiography; ED, emergency department; OR, odds ratio; PE, pulmonary embolism; RV, right ventricular.

### Agreement with manual review

Agreement on the four-category statement in the development sample increased from 113/150 (75.3%, κ 0.67) before revision to 133/150 (88.7%, κ 0.85) after tuning on the same sample. In the held-out sample, the rules agreed with manual physician coding of the four-category overall strain statement in 93 of 100 reports (κ 0.89, 95% CI 0.81-0.97; Tables 5 and 6). Sensitivity for detecting any overall statement was 97.4% (74/76, 95% CI 90.9-99.3%) and specificity was 91.7% (22/24, 95% CI 74.2-97.7%). This agreement applies only to the classification of overall strain statements. The held-out sample contained no numerical RV/LV ratios, so the ratio estimates rest on the cohort-wide coding rules and supplementary search, with manual confirmation of every flagged report; the sensitivity of this approach for unusual ratio wording could not be estimated.

**Table 5.** Agreement between rule-based coding and manual physician coding.

| **Variable** | **Agreement (95% CI)** | **Kappa (95% CI)** |
|---|---|---|
| Overall RV strain statement (4 categories) | 93.0% (86.3–96.6) | 0.89 (0.81–0.97) |
| Interventricular septum mentioned | 100% (96.3–100) | 1.00 |
| IVC reflux mentioned | 99.0% (94.6–99.8) | 0.90 (0.65–1.00) |
| RV size mentioned | 97.0% (91.5–99.0) | 0.89 (0.74–1.00) |
| Qualitative RV–LV size comparison | 95.0% (88.8–97.8) | 0.43 (0.00–0.80) |
| RV/LV ratio | No ratios in sample | Not estimable |

CI, confidence interval; IVC, inferior vena cava; RV, right ventricle; LV, left ventricle.

**Table 6.** Classification of overall RV strain statements by the physician and rule-based coding.

| **Physician category** | **Absent** | **Hedged** | **Present** | **No statement** | **Total** |
|---|---|---|---|---|---|
| Absent | 48 | 0 | 1 | 1 | 50 |
| Hedged | 0 | 17 | 2 | 1 | 20 |
| Present | 0 | 0 | 6 | 0 | 6 |
| No statement | 2 | 0 | 0 | 22 | 24 |
| Total | 50 | 17 | 9 | 24 | 100 |

Rows show the physician’s classifications and columns show the rule-based classifications. RV, right ventricular.

## Discussion

Exact RV/LV ratios were recorded in five of 1,534 positive CTPA reports, and only seven additional reports recorded a numerical threshold or range. Of the 412 reports without an overall strain statement, 39 still described at least one relevant CT feature. The proportion of reports lacking RV documentation therefore depends on whether the audit counts only overall interpretations or also credits descriptions of individual features. Under the broader definition, the proportion of reports containing neither an overall RV strain statement nor any of the four coded CT features was 24.3% (373 reports).

The distinction between an exact ratio and a threshold matters when assessing uptake of the 2026 recommendation [4]. A statement such as “RV/LV ratio greater than 1” conveys that a cutoff was exceeded. The measured value also conveys the magnitude of relative RV enlargement. An exact-ratio field alongside the overall interpretation would allow reports to communicate both quantitative information and the radiologist’s assessment, including uncertainty when appropriate. Our data establish how often these elements appeared in historical reports. The underlying measurements and their use in clinical risk stratification could not be assessed.

Earlier CTPA audits documented variability in how radiologists reported RV strain [5,6]. The present study extends those observations by quantifying exact ratios, thresholds, and qualitative descriptions separately and by examining documented CT features across all four categories of overall strain statements. This distinction between the finding and the way it is documented also applies to other cardiovascular findings visible on CTPA, including coronary artery calcification, which has been reviewed as a potential prognostic marker in acute PE [15].

Omission of an overall strain statement became markedly less frequent during the study period. After adjustment for examination period, isolated subsegmental PE remained associated with higher odds, and ED linkage with lower odds, of both types of documentation gap. The association between the structured heading and omission attenuated after adjustment for estimated examination period. The heading was absent in the two earliest periods, limiting our ability to distinguish its association from changes in reporting practice over time. These findings identify reporting contexts that could be examined when introducing a dedicated RV/LV field. A prospective audit could track exact-ratio documentation and record when technical limitations prevent measurement.

## Limitations

This retrospective analysis included the first dated positive CTPA report per patient at a single center. The absence of images precluded verification of ventricular measurements and assessment of technical factors that might have limited measurement. The study also could not assess whether documentation influenced care. One physician performed all manual coding, so inter-reader agreement could not be estimated. The held-out sample contained no numerical ratios, and agreement for qualitative RV-LV comparisons was lower than for overall strain statements. The numerical-ratio estimate depended on the coding rules and the supplementary search, followed by manual verification of flagged reports. Their sensitivity for unconventional ratio wording is unknown, so some ratios may have been missed. Because dates were shifted, examination years could be estimated only to within 1 year, so adjustment for examination period was approximate. Report format and examination period were closely linked, so their separate contributions could not be distinguished. The estimates describe historical local practice and may differ from contemporary practice at other centers.

## Conclusion

In 1,534 positive CTPA reports, 0.3% documented the exact RV/LV ratio and 24.3% documented neither an overall RV strain statement nor any of the four CT features assessed. Omission of an overall statement declined over time. These findings provide a single-center baseline for evaluating implementation of the 2026 numerical-ratio reporting recommendation.

## Data Availability

The de-identified data used in this study are available through PhysioNet to credentialed users who complete the required training and sign the applicable Data Use Agreements. The data sources were MIMIC-IV-Ext-PE v1.0.0 (https://www.physionet.org/content/mimic-iv-ext-pe/1.0.0/), MIMIC-IV v3.1 (https://physionet.org/content/mimiciv/3.1/), and MIMIC-IV-Note v2.2 (https://www.physionet.org/content/mimic-iv-note/2.2/).

https://www.physionet.org/content/mimic-iv-note/2.2/

https://physionet.org/content/mimiciv/3.1/

## Statements and declarations

### Ethics approval and consent

The institutional review boards at Beth Israel Deaconess Medical Center and the Massachusetts Institute of Technology approved creation of the de-identified MIMIC databases with a waiver of individual consent. The secondary analysis used credentialed access under the PhysioNet data use agreement. No ethics approval was required for this secondary analysis.

### Data and code availability

MIMIC-IV-Ext-PE, MIMIC-IV, and MIMIC-IV-Note are available to credentialed users through PhysioNet. Analysis code and coding rules are available under the MIT License. The repository URL is withheld for double-anonymous review.

### Use of artificial intelligence

No AI was used.

